# A survey of clinical opinions and preferences on the non-surgical management of intermittent exotropia in China

**DOI:** 10.1101/2021.04.13.21255451

**Authors:** Yidong Wu, Tingting Peng, Jinjing Zhou, Meiping Xu, Yi Gao, Jiawei Zhou, Fang Hou, Xinping Yu

## Abstract

**Purpose:** Intermittent exotropia (IXT) is the most common form of childhood exotropia. Currently, controversies exist regarding its management and non-surgical options in particular. This study reports clinical opinions and preferences on the non-surgical management among practitioners in China. The opinions within and between ophthalmologists and optometrists were also compared.

**Design:** Cross-sectional survey study.

**Methods:** An online survey was developed and distributed through professional bodies. The study was conducted from July 25^th^ to August 3^rd^, 2019. A total of 300 ophthalmologists and 188 optometrists responded.

**Results:** Of 488 participants, 257 (53%) considered fusion defects as the main cause of IXT, and 299 (61%) took IXT as a progressive disorder. Two hundred and seventy-one (56%) participants considered orthoptic exercises as the most effective non-surgical intervention for IXT. Likewise, 245 (50%) participants reported that orthoptic exercises was their most frequent non-surgical option, followed by observation (178, 37%). There are discrepancies between ophthalmologists and optometrists. A greater proportion of ophthalmologists (201, 67%) shared the view that IXT worsens over time compared with optometrists (98, 52%) (*p* = 0.001). Additionally, ophthalmologists (121, 40%) tended to prefer observation compared with optometrists (57, 30%) (*p* = 0.021).

**Conclusions:** This study shows that there is no general consensus on the non-surgical management of IXT in China. Given the lack of robust evidence, the findings from this study not only show the current clinical opinions but also highlight the need for future randomized clinical trials to validate the effectiveness of non-surgical interventions, orthoptic exercises in particular, and to establish treatment guidelines accordingly.

## Introduction

Intermittent exotropia (IXT) is the most common form of childhood strabismus in Asia^1, 2^, with an incidence rate of approximately 4% in China^3, 4^. If untreated, IXT may lead to loss of binocular vision and can induce psychosocial problems^5-7^. Conventionally, surgery is often the choice of intervention when there is poor control of an exodeviation^8, 9^. However, the surgical approach comes with a risk of persistent overcorrection (esotropia), which may result in diplopia, development of amblyopia, and reduction in stereopsis^8^. There is also evidence that the surgical benefits decline over time, with a strong tendency for recurrence^10-12^. In light of these concerns, non-surgical management, due to its noninvasive nature, has been recommended to defer or even avoid surgery^13, 14^. Current non-surgical options include observation^15^, part-time patching^16^, overminus lenses^17^, prisms^18^, and orthoptic exercises^19^. However, to date, there is not a clear understanding of which option benefits patients most. The optimal timing of interventions, the dose-response relationship and the long-term efficacy are also not clearly understood^8, 13^. Consequently, there is a dispute regarding the non-surgical management of IXT.

Three reasons motivated us to conduct this survey. First, the prevalence of IXT is dramatically higher in China than that in the Western countries (nearly 4:1)^3, 4^. This means that a greater number of people need medical care for IXT in China. It is, therefore, essential to clarify the current clinical opinion. Second, there are no official guidelines regarding the management of IXT in China. The choice of treatment option is often guided by practitioners’ previously held beliefs. Third, unlike the medical education system in the West, Chinese optometric education is also medically-based^20-22^. After gaining medical licenses, practitioners are assigned to perform as optometrists, specializing in refractive correction and primary eye care^20, 21^. Typically, the surgical management of IXT is conducted by ophthalmologists, and the non-surgical management of IXT is undertaken by both ophthalmologists and optometrists. We are interested in whether there are discrepancies in the management of IXT between these two groups of professionals.

We are not aware of any reports regarding the clinical opinions and preference on non-surgical management of IXT in China. Here, we conducted a survey to shed light on this topic. The opinions within and between ophthalmologists and optometrists were also compared.

## Methods

The study conformed to the Declaration of Helsinki and was approved by the Ethics Committee of the Eye Hospital of Wenzhou Medical University.

The initial survey questions were identified during discussions with an expert group consisting of three ophthalmologists and two optometrists. Subsequently, the questionnaire was validated with 20 practitioners at our institutes. Based on their responses and comments, the questions were amended. The final survey questions (Table 1) can be grouped into (1) professional role and characteristics (questions 1 - 4) and (2) clinical opinions and preferences (questions 5 - 9). The survey was made available online through *Wenjuanxing (www.wjx.cn)*. The questionnaires were distributed through professional bodies (Chinese Association for Pediatric Ophthalmology and Strabismus, National Ophthalmology & Optometry Alliance), personal contacts, and social media (WeChat) between July 25^th^, 2019, and August 3^rd^, 2019. Participants were not allowed to skip questions (i.e., each question had to be answered), while for some questions, an “other” option was available if the provided options did not match their policies. Participants were informed about the purpose of the survey and voluntary participation. Data were collected anonymously and did not contain identifying information. For statistical analysis, the Chi-square test and Fisher’s exact test were used with an alpha value of 0.05 in SPSS, version 25.0 (SPSS, Inc., Chicago, IL, USA), and Bonferroni correction was applied if multiple comparisons were performed.

**Table 1.** Survey questions

| Survey questions | Response options |
| --- | --- |
| 1. Which of the following best describes your current occupation? | Ophthalmologist<br>Optometrist |
| 2. How many years have you been in practice? | 1~5<br>6~10<br>11~15<br>> 15 |
| 3. How many patients with IXT do you examine in a typical month? | < 5<br>6~10<br>11~20<br>21~49<br>50~100<br>> 100 |
| 4. Which age group of patients with IXT do you examine most frequently? | < 4<br>5~12<br>13~18<br>> 18 |
| 5. In your opinion, what might be the main cause of IXT? | Abnormality of extraocular muscles<br>Orbital factors<br>Defects of fusion mechanisms<br>Defects of other cortical mechanisms<br>Uncertain<br>'Other' with an open-ended response |
| 6. In your opinion, what is the natural course of IXT for the majority of patients (in terms of the magnitude of exodeviation, control of exodeviation and (or) stereoacuity)? | Progressive |
|  | Stable |
|  | Improved |
|  | Each of the above is possible |
| 7. Which of the following non-surgical interventions do you consider most effective for the majority of patients with IXT? | Overminus lenses |
|  | Prisms |
|  | Part-time patching |
|  | Orthoptic exercises |
|  | Botulinum toxin A injection |
|  | All of the above are ineffective |
|  | Uncertain |
| 8. In your practice, which of the following is your most frequent non-surgical option other than the correction of refractive error? | Overminus lenses |
|  | Prisms |
|  | Part-time patching |
|  | orthoptic exercises |
|  | Botulinum toxin A injection |
|  | Observation |
|  | 'Other' with an open-ended response |
| 9. Do you agree with the following opinions regarding early surgery for IXT to gain superior sensory outcomes? |  |
| Surgery at a younger age | Yes/No/Uncertain |
| Surgery within the critical period | Yes/No/Uncertain |
| Surgery after a shorter duration of strabismus | Yes/No/Uncertain |
| Surgery when the severity of IXT increases (angle of deviation, control, stereoacuity) | Yes/No/Uncertain |
| IXT = intermittent exotropia |  |

## Results

A total of 488 practitioners responded. Because no report is available regarding how many practitioners in China provide strabismus management services, we cannot calculate exact response rate. According to the electronic database powered by *http://www.haodf.com* (the largest online healthcare community in China), there were 1251 active practitioners providing eye care services concerning strabismus at the time of the survey. Therefore, we achieved an estimated response rate of 39%.

### Participants’ profile

The participants included 300 ophthalmologists and 188 optometrists. Of the 300 ophthalmologists, 206 (69%) had been in practice for more than ten years. Of the 188 optometrists, 96 (51%) had been in practice for more than ten years. One hundred and seventy-seven (59%) ophthalmologists reported that they examined more than ten patients per month, whereas 143 (76%) optometrists reported examining fewer than ten patients per month. Most participants (392, 80%) of both professionals reported that the age group of their patients was between 5∼12 years old.

### Clinical impression

#### Possible Etiology

The factors that practitioners considered as the main cause of IXT are shown in Table 2. Of 488 participants, 257 (53%) considered fusion defects as the main cause of IXT, followed by abnormality of extraocular muscles (117, 24%), deficits of other cortical mechanisms (44, 9%) and orbital factors (1, 1%). Besides, 60 (12%) participants were uncertain about the main cause of IXT. Of nine participants who chose the option ‘Other’, five left specific responses: four gave answers in relation to refraction; one argued that IXT was a multifactorial disease without a primary cause. There was no significant difference between ophthalmologists and optometrists (Fisher’s exact test, *p* = 0.214).

**Table 2.** The factors that practitioners considered as the main cause of intermittent exotropia.

| Factors | Ophthalmologists*, No. (%) | Optometrists*, No. (%) | All, No. (%) |
| --- | --- | --- | --- |
| Abnormality of extraocular muscles | 68 (23) | 49 (26) | 117 (24) |
| Orbital factors | 0 (0) | 1 (1) | 1 (1) |
| Defects of fusion mechanisms | **160 (53)** | **97 (52)** | **257 (53)** |
| Defects of other cortical mechanisms | 33 (11) | 11 (6) | 44 (9) |
| Uncertain | 33 (11) | 27 (14) | 60 (12) |
| Other | 6 (2) | 3 (2) | 9 (2) |
| Total | 300 (100) | 188 (100) | 488 (100) |

The results in bold show the most frequent response. \*Fisher exact test, $p = 0.214$ .

#### Natural course

As shown in Table 3, 299 (61%) participants considered IXT as a progressive disorder. Only a small minority (5, 1%) held the view that IXT might improve over time in terms of the magnitude of exodeviation, control of exodeviation and (or) stereoacuity. A Fisher’s exact test indicated a significant difference between ophthalmologists and optometrists (*p* = 0.009). Compared with optometrists (98, 52%), a greater proportion of ophthalmologists (201, 67%) shared the view that IXT worsens over time (Chi-square test, χ^2^ = 10.77, *p* = 0.001).

**Table 3.**
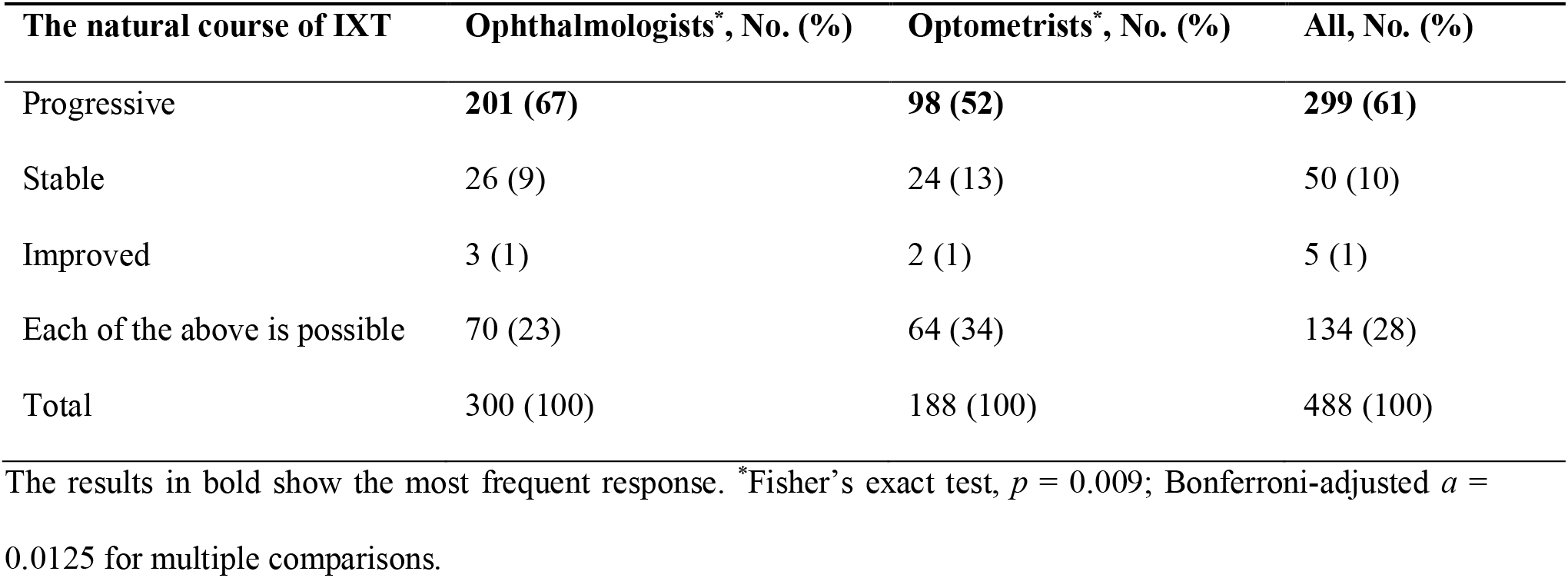
The clinical impression of the natural course of intermittent exotropia.

### Non-surgical management

#### Effectiveness of non-surgical interventions

Among all the non-surgical interventions, orthoptic exercises were considered the most effective by both professionals (ophthalmologists, 161, 54%; optometrists, 110, 59%; Table 4). The choices of other interventions, including overminus lenses (6%), prisms (11%), part-time patching (2%), and botulinum toxin A injection (3%), showed a scattered pattern. Notably, 64 (13%) participants indicated that all non-surgical interventions were ineffective, and 47 (10%) participants claimed that they were uncertain about which one works more effectively. A Fisher’s exact test indicated that opinions differed significantly between ophthalmologists and optometrists (*p* = 0.001). Multiple comparisons found that (i) a greater proportion (32, 17%) of optometrists considered prisms as an effective intervention than that of ophthalmologists (22, 7%) (Chi-square test, χ^2^ = 11.11, *p* = 0.001), and that (ii) ophthalmologists (50, 17%) were more likely to feel that all these non-surgical interventions were ineffective compared with optometrists (14, 7%) (Chi-square test, χ^2^ = 8.62, *p* = 0.003).

**Table 4.** The practitioners’ opinions on the effectiveness of non-surgical interventions.

| Interventions | Ophthalmologists*, No. (%) | Optometrists*, No. (%) | All, No. (%) |
| --- | --- | --- | --- |
| Overminus lenses | 21 (7) | 7 (4) | 28 (6) |
| Prisms | 22 (7) | 32 (17) | 54 (11) |
| Part-time patching | 3 (1) | 5 (3) | 8 (2) |
| Orthoptic exercises | **161 (54)** | **110 (59)** | **271 (56)** |
| Botulinum toxin A injection | 12 (4) | 4 (2) | 16 (3) |
| All of the above are ineffective | 50 (17) | 14 (7) | 64 (13) |
| Uncertain | 31 (10) | 16 (9) | 47 (10) |
| Total | 300 (100) | 188 (100) | 488 (100) |

The results in bold show the most frequent response. \*Fisher's exact test, $p = 0.001$ ; Bonferroni-adjusted $\alpha =$
0.007 for multiple comparisons.

#### The most frequent non-surgical management option

Subsequently, the practitioners were asked the most frequently applied non-surgical option in their practice other than the correction of refractive error. Likewise, both professionals reported orthoptic exercises as the most frequent option (ophthalmologists, 144, 48%; optometrists, 101, 54%), followed by observation (ophthalmologists, 121, 40%; optometrists, 57, 30%; Table 5). Apart from orthoptic exercises and observation, all other management options were rarely chosen (sum = 13%, 65/488). Two ophthalmologists chose “Other” and left detailed responses. Their responses were similar to observations (i.e., do not take any specific intervention unless the surgery was needed). Therefore, we merged them into the category “Observation” for analyses. A Fisher’s exact test indicated a significant difference between ophthalmologists and optometrists (*p* = 0.024). We then conducted multiple comparisons. Compared with optometrists (57, 30%), ophthalmologists (121, 40%) tended to prefer observation. (Chi-square test, χ^2^ = 8.62, *p* = 0.021, *a* = 0.008).

**Table 5.** The practitioners’ most frequent non-surgical options for patients with intermittent exotropia.

| Interventions | Ophthalmologists*, No. (%) | Optometrists*, No. (%) | All, No. (%) |
| --- | --- | --- | --- |
| Overminus lenses | 15 (5) | 7 (4) | 22 (5) |
| Prisms | 15 (5) | 20 (11) | 35 (7) |
| Part-time patching | 2 (1) | 3 (2) | 5 (1) |
| Orthoptic exercises | **144 (48)** | **101 (54)** | **245 (50)** |
| Botulinum toxin A injection | 1 (1) | 0 (0) | 1 (1) |
| Observation | 121 (40) | 57 (30) | 178 (37) |
| Other | 2 (1) | 0 (0) | 2 (1) |
| Total | 300 (100) | 188 (100) | 488 (100) |

The results in bold show the most frequent response. \*Fisher’s exact test, $p = 0.024$ ; Bonferroni-adjusted $\alpha = 0.008$ for multiple comparisons.

#### Early surgery to gain superior sensory outcomes

In the last question, the participants were asked whether they agreed with the idea regarding early surgery to gain superior sensory outcomes of four perspectives: (i) surgery at a younger age; (ii) surgery within the critical period; (iii) surgery after a shorter duration of strabismus; and (iv) surgery while the severity of IXT increases in terms of either angle of exodeviation or control of exodeviation or stereoacuity. Their responses are shown in Table 6. Most participants (449, 92%) agreed with perspective (iv), whereas opinions varied on perspectives (i) ∼ (iii). Compared with the optometrists, fewer ophthalmologists agreed with the idea from perspectives (i) ∼ (iii) (Chi-square test, χ^2^ = 8.919, *p* = 0.003; χ^2^ = 6.936, *p* = 0.009; χ^2^ = 10.794, *p* = 0.001; respectively).

**Table 6.**
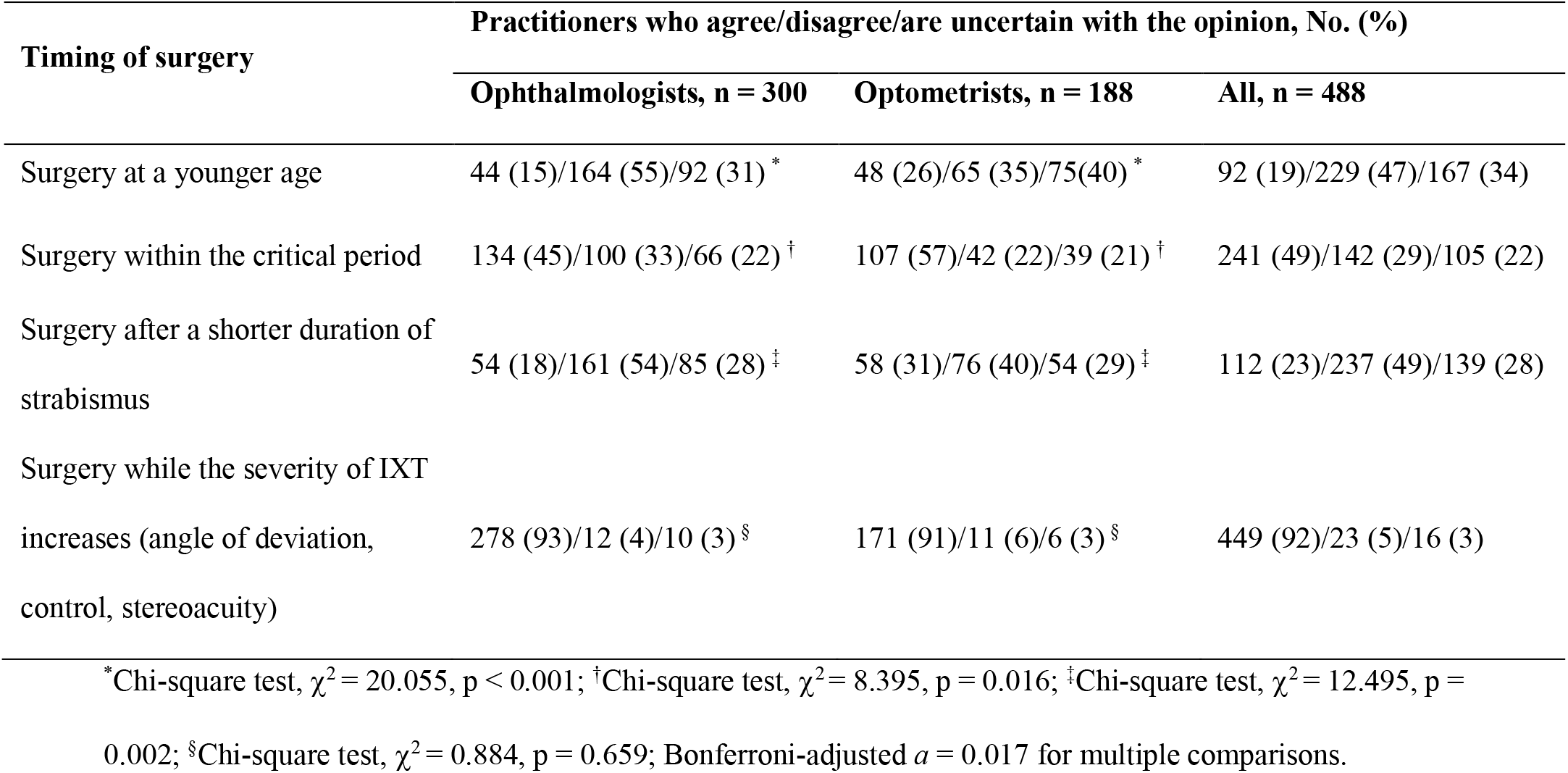
The practitioners’ opinions regarding early surgery for intermittent exotropia.

### Additional analysis

Whether a relationship between the clinical impression (Table 2, 3) and the choice of management option (Table 5) is of great interest. Since a vast majority of participants reported orthoptic exercises (50%) and observation (37%) as their most frequent management option (Table 5), we included these two options for further investigation. We tested two plausible hypotheses: (i) Participants who considering IXT as a disorder mainly caused by fusion defects would prefer orthoptic exercises than other management options and (ii) Participants who holding the view that IXT worsens over time were more prone to intervention rather than observation.

Among 257 participants who taking fusion defects as the main cause of IXT, 142 (55%) reported orthoptic exercises as the most frequent non-surgical management option. This rate was significantly higher than that (103, 45%) of participants who considering other factors as the main cause of IXT (Chi-square test, χ^2^ = 5.534, *p* = 0.019).

Of 299 participants who taking IXT as a progressive disorder, 113 (38%) reported observation as their most frequent option. This rate was not significantly different from that (67, 35%) of participants who holding other views on the natural course of IXT (Chi-square test, χ^2^ = 0.273, *p* = 0.601).

## Discussion

In this survey study, we aimed to show the opinions of practitioners rather than to provide a Preferred Practice Pattern or an Expert Consensus. Our results demonstrated both consistencies and discrepancies regarding the non-surgical management of IXT among practitioners in China. We showed that both ophthalmologists and optometrists considered orthoptic exercises as most effective non-surgical intervention. Likewise, both professionals chose orthoptic exercises as their most frequent non-surgical management option, followed by observation. We found that a greater proportion of ophthalmologists shared the view that IXT worsens over time and tended to prefer observation.

The preference for orthoptic exercises may reflect the reality in a broader Asian area. In a Hong Kong cohort of 117 children with IXT, Kwok et al. reported that orthoptic exercise was taught to all of their patients^23^. In an Israeli survey, 66% (37) of the respondents reported that they used orthoptic exercises for IXT treatment, a percentage higher than that of respondents using part-time patching (52%, 29) and overminus lenses (48%, 27)^24^.

It is likely that our findings, especially the preference for orthoptic exercises, are different from the ophthalmologists’ practice pattern in Western countries. In 1990, a survey of members of the American Association for Pediatric Ophthalmology and Strabismus (AAPOS) showed that 31 (48%) of the respondents rarely or never used non-surgical interventions^25^. Of the 34 (52%) respondents who routinely used one or more non-surgical techniques, a majority chose part-time patching (46%) and minus lenses (34%) as the most frequent interventions. In contrast, orthoptic exercises were less frequently used (14%)^25^. In a cohort study of 460 children with IXT between May 2005 and December 2006 in the United Kingdom, Buck and her colleagues reported that 65% (297) of the children had no specific treatment (i.e., observation) within the first year of diagnosis^26^. Among the 36 (8%) children who received non-surgical interventions other than correction of refractive error, the treatment methods were overminus lenses (21), part-time patching (10) and orthoptic exercises (5)^26^. The aforementioned two studies indicated that observation, part-time patching and overminus lenses were more popular non-surgical management options than orthoptic exercises in Western countries. Indeed, it has been suggested that orthoptic exercises in the management of IXT had been on the decline^13, 27, 28^. This decline might be due to the surgeons’ critical attitude towards the role of orthoptic exercises, namely, it has not been proven that orthoptic exercises would provide a benefit when combined with surgeries, or that orthoptic exercises could replace surgeries^13, 28, 29^.

Our survey cannot directly reveal the underlying reasons for the preference for orthoptic exercises. Why do our participants prefer orthoptic exercises rather than other management options? Two possible explanations are discussed here. First, the choice of orthoptic exercises is consistent with the participants’ held beliefs. As shown in Table 2, most practitioners considered IXT to be a disorder caused by fusion defects (53%). Theoretically, orthoptic exercises increase fusional vergence ranges and normalize sensory fusion, thereby facilitating exotropia control^13^. Our *Additional analysis* further supports this explanation. Second, we speculate that a historical reason might contributed to the preference for orthoptic exercises. Optometry, as an emerging discipline in China since the 1980s, was established by Chinese ophthalmologists with support from American optometrists^20-22^. This enables a unique cooperation between ophthalmology and optometry in China. It has been suggested that orthoptic exercises are favored by American optometrists in the management of IXT^28^. In Rutstein and Corliss’ cohort of 73 patients with IXT, 82% (60/73) of patients had non-surgical treatment, with 49% (36/73) having orthoptic exercises^30^.

Overall, the indication and effectiveness of non-surgical interventions have not been established^8, 13^. Most published reports comprised retrospective studies from single institutions. In Coffey et al.’s general review of treatments for IXT, the pooled success rates were as follows: overminus lenses (n = 215), 28%; prism (n=201), 28%; part-time patching (n=170), 37%; orthoptic exercises (n=740), 59%; surgery (n=2530), 46%^31^. Caution should be taken when interpreting these results because there are great variations in the criteria for “success” among the authors (e.g., follow-up periods, the magnitude of deviation), as well as in the sample selection criteria. Recently, the Pediatric Eye Disease Investigator Group (PEDIG) conducted a series of randomized clinical trials to validate the effectiveness of non-surgical management options, including part-time patching^16^, overminus lenses^17^ and observation^15^. PEDIG found that patients who received non-surgical interventions, either part-time patching^16^ or overminus lenses^17^, gained slight benefits in a short-term period (six months for part-time patching, eight weeks for overminus lenses). However, they also found that deterioration was uncommon over a 3-year observational period in 3- to 10-year-old children with IXT^15^. Put differently, it’s unconvincing to recommend one treatment option over the others if the treatment rationale for IXT is “preventing deterioration”.

In the survey, we included a question regarding surgical indication (Table 6). The reason is that surgical indication is a critical topic during the non-surgical management and may help to explain the non-surgical practice pattern. Since there is a long-existing debate concerning whether earlier surgery can achieve superior sensory outcomes for patients with IXT^32-36^, we narrowed our question into this scope of four perspectives (see Table 5 for details). Only a minority of participants agree with the view that surgery at a younger age (19%) or after a shorter duration (23%) might achieve superior sensory outcomes. These findings suggested that most of the practitioners were likely to apply non-surgical option first when managing their patients. Therefore, non-surgical procedure is a large component in the management of IXT.

Our study has limitations. First, we could not calculate the exact response rate due to the recruitment method. There was also no previously published survey in this population that we could use for comparison. However, this study is likely to be a good representation because we were privileged to contact as many practitioners as possible with support from the National Clinical Research Center for Ocular Diseases and several professional bodies. Second, like all survey-based analyses, our results represented opinions, which might not necessarily agree with actual behaviors. Third, our study only questioned the “most frequency” option, which limited our ability to determine the frequency of each management option. This choice of question form was because we wanted to make the questionnaire brief so that more practitioners might respond. In addition, the discrepancies between ophthalmologists and optometrists revealed in this study should be interpreted with caution. It is possible that the patients who turned to ophthalmologists were severer cases.

In conclusion, our study showed that although a majority of practitioners reported orthoptic exercises as most frequent non-surgical option, there was no general consensus among practitioners concerning the non-surgical management of IXT in China. Given the high prevalence of IXT in the Asian population^1-4^ and the lack of robust evidence, our findings emphasize the need for future randomized controlled trials in Asian area to determine the effectiveness of non-surgical interventions, and to establish treatment guidelines for IXT accordingly.

## Data Availability

The data used to support the findings of this study are available from the corresponding author (XY) upon request request.

## Acknowledgments

We would like to thank all the participants who took part in the survey. We are also grateful to the following professional bodies for their kind assistance: Chinese Association for Pediatric Ophthalmology and Strabismus, National Ophthalmology & Optometry Alliance.

